# The Experience of Automated Home Oxygen Therapy for Patients with COPD – a Qualitative Study

**DOI:** 10.1101/2024.11.02.24316557

**Authors:** Linette Marie Kofod, Louise Bolvig Laursen, Elisabeth Westerdahl, Ejvind Frausing Hansen, Barbara Cristina Brocki, Morten Tange Kristensen, Dorthe Gaby Bove

## Abstract

The present study included the first patients with COPD on long-term oxygen therapy who experienced second-by-second oxygen adjustments in their homes based on oxygen saturation. A device capable of automatically titrating the patient’s oxygen was installed in the patients’ home aiming at increasing the time spent within target saturation. We explored patients’ experiences with this automated home oxygen titration, focusing on how maintaining target saturation affected daily life.

Semi-structured interviews were conducted with eight men and four women after installation. Systematic text condensation was used in the analysis.

Three main themes emerged from patient narratives: 1) “This is my life” – Patients preferred maintaining stable oxygen saturation, associating hypoxemia with dyspnea, discomfort, and difficulties with daily tasks. 2) “Getting the oxygen, I need” – Many patients reported improved ability to perform daily activities when oxygen was automatically adjusted. 3) “New technology gives hope for my life” – Patients expressed optimism about the potential of home-based technology, offering suggestions to improve usability, mainly by reducing concentrator noise.

Our findings suggested high acceptability of the automated oxygen in the patients’ home, as they believed it to increase the time spend with sufficient oxygen, especially during daily activities. Integrating patient insights is essential for implementation and acceptance of automated home oxygen therapy.

## Introduction

The technological development is advancing in the area of long-term oxygen therapy (LTOT), leading to options for optimizing treatment for patients in need of this treatment. Especially people with severe Chronic Obstructive Pulmonary Disease (COPD) can develop a chronic respiratory failure leading to the need of LTOT. The quality of life in this patient group is often poor, characterized by dyspnea, low activity levels, social isolation and high mortality [1-3]. It has long been recognized that despite being prescribed a fixed oxygen flow at rest, patients with chronic respiratory failure frequently experience drops in oxygen saturation when engaging in physical activity [4-6]. Traditionally, these desaturation episodes have been tolerated by the healthcare professionals without adjustments of the prescribed oxygen flow [7,8]. As long as the patients’ oxygen saturation is kept within a target range of 90-94% at rest, interventions for hypoxemia during physical activities at home have been limited.

Innovations in the field have led to the development of closed-loop oxygen delivery devices capable of automated oxygen titration based on real-time oxygen saturation measurements. This technology has demonstrated the ability to minimize desaturation episodes by automatically adjusting the oxygen flow to meet the patients’ immediate needs. Both during hospitalization and in walking tests, automated oxygen titration increased the time the patients spent within the target saturation range, resulting in less dyspnea and longer walking distances [9-13]. Furthermore, a crossover study showed that the patients appeared to benefit from automated oxygen adjustments during an activity of daily living (ADL) test [14]. The patients not only enhanced their performance in the ADL-test, but they also reported less dyspnea when oxygen saturation was kept more within target. These findings suggest that patients may benefit from optimal oxygenation during everyday tasks. Consequently, we conducted a pilot study in which the closed-loop device was installed in the homes of 12 patients with COPD already on LTOT, aiming to assess the feasibility of improving oxygenation through automated titration. As the technology has now been adapted for home oxygen use, it is essential to consider patients’ perspectives and attitudes toward the development of having an automated system managing their oxygen therapy at home.

Therefore, this study aimed to explore the experiences of patients receiving long-term oxygen therapy (LTOT) at home using automated oxygen titration, with a particular focus on how it affected their daily lives.

## Materials and methods

### Design

This qualitative study used interviews to explore the experiences of patients treated with automated oxygen administration at home. The study was nested in the pilot randomized crossover feasibility trial titled: Automatically adjusted oxygen in response to saturation during daily life of patients with COPD on long-term oxygen therapy (DaiLiHOT_2), ClinicalTrials.gov no. NCT05556187.

The study was based on systematic text condensation as describe by Malterud,[15] and complied with the Consolidated Criteria for Reporting Qualitative Research (COREQ) [16].

### Setting and participants

The study participants were recruited from the Departments of Pulmonology at Copenhagen University Hospital, Hvidovre and Copenhagen University Hospital, Bispebjerg-Frederiksberg, Denmark as a part of the DaiLiHOT_2 study (in draft).

A full description of recruiting and intervention in presented in the abovementioned DaiLiHOT_2 study. Briefly, all included participants were diagnosed with COPD and received long term oxygen treatment. The inclusion criteria were chronic respiratory failure with resting hypoxemia (PaO2 ≤ 7.3 kPa). The patients should be able to walk independently, with or without walking aid, cognitively able to participate and provide informed consent. Patients received four days of oxygen treatment delivered by a continuous automated titration of the oxygen flow based on the patients’ saturation using the electronic closed-loop device O2matic HOT (O2matic Ltd., Herlev, Denmark) installed in their home. The aim was to keep the saturation within a target from 90-94% by letting the oxygen flow vary from 0.9-6.6 L/min according to the algorithm in the device. The patients’ normal 5 L/min oxygen concentrator was upgraded to a 9 L/min concentrator. The patients wore a Wrist Pulse Oximeter (Nonin Medical, Inc., USA), which transmitted data on oxygen saturation to the HOT device using Bluetooth. Further, the patients received a tablet on which they could follow the treatment. The setup is shown in Figure 1. When leaving their homes, the patients used their regular portable oxygen concentrator with a fixed oxygen flow.

**Figure 1.**
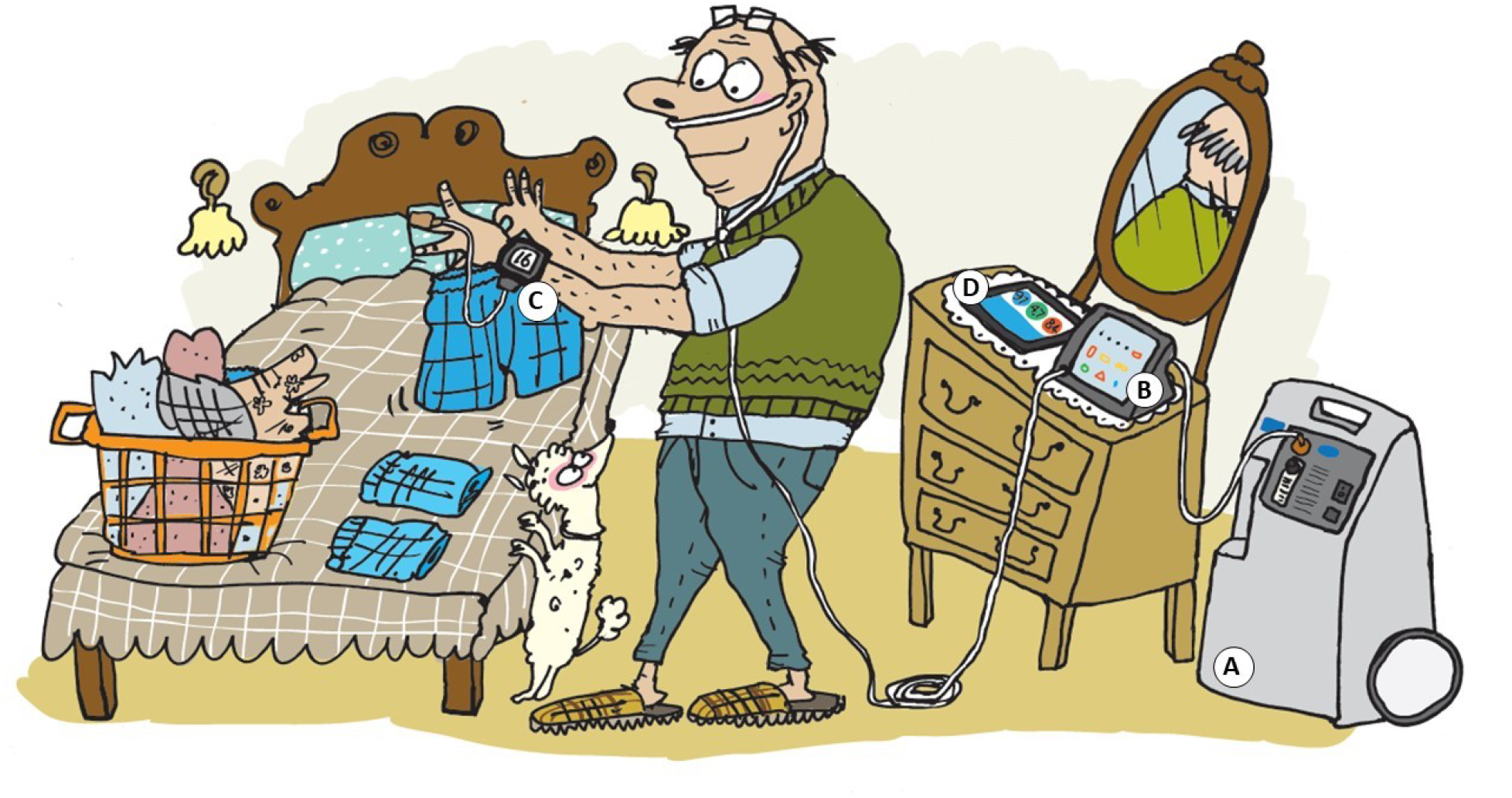
Artistic presentation of the setup of automated oxygen titration in the patient’s home. (A) Oxygen concentrator, (B) Automated oxygen titration device, (C) Wrist pulse oximeter, (D) Patient tablet

A total of 12 patients were treated with automated oxygen adjustments at home (DaiLiHOT_2 study) and all were invited to participate in this interview study.

### Data collection

The interviews, besides one, were conducted in the patients’ homes from January to December 2023. One interview was conducted in a meeting room at the hospital according to patient preference.

The majority of the interviews were conducted by the second author (n=9), and a few by the first author (n =3). The first author is an experienced respiratory physiotherapist and part of the patients’ treatment team. The second author is a trained interviewer and not part of the patients’ treatment team and did not know the patients beforehand. All interviews were recorded and transcribed verbatim by the first author.

The semi-structured interview guide shown in Table 1 was based on discussions among the authors and existing literature. The first question aimed to gather an impression of the patients’ everyday life with usual care, providing the interviewer with a starting point. The interview technique was open-ended, with follow-up questions based on the patients’ responses or the level of detail they provided initially. Probes such as “Can you explain in more detail?” or “Can you give an example?” were used to deepen and nuance the responses. If a spouse was present during the interview, they were invited to elaborate on or further clarify the patient’s statements.

**Table 1.**
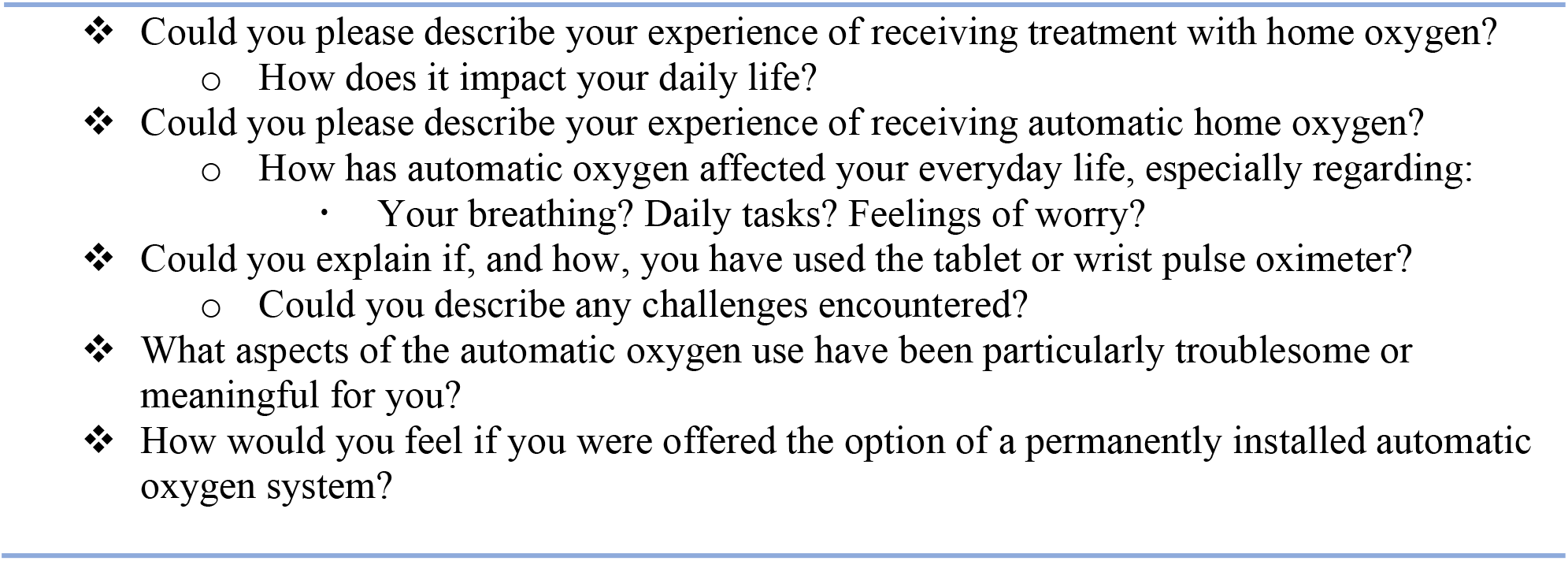
Opening questions for the semi-structured interviews.

Everything was transcribed; however, the spouses’ contributions were not included in the analysis.

### Data analysis

The data analysis was conducted inductively and followed the systematic text condensation, as outlined by Malterud [15]. Initially, the first and second authors independently read and re-read the transcripts multiple times to gain a comprehensive understanding and form an initial impression of the content. Following this, the material was systematically coded. The first and second author performed coding independently, after which their individual codes were compared and discussed. Where discrepancies arose, they were resolved by merging the differing codes into new, agreed-upon codes. Subsequently, ‘Meaning Units’ were identified, which were then grouped into preliminary categories or subthemes. These subthemes were subjected to multiple rounds of discussion among the authors, leading to the resolution of differences and the establishment of consensus. This process facilitated the development of tentative, superior themes, which were further refined and rewritten through additional discussions until coherent themes and subthemes were achieved. Patient quotes were anonymized with made up names. As a final step, the original transcripts were revisited to ensure that the results accurately reflected the data and that the “patients’ voices” were authentically represented.

Instead of employing qualitative data analysis software, we maintained rigorous data organization manually, utilizing paper, post-it notes, and a large whiteboard to visually manage and structure the data.

### Ethical considerations

All participants were informed both in writing and verbally about the aim of the study and its voluntary nature. They were given a minimum 24 h to consider their participation and the opportunity to ask detailed and clarifying questions. All participants signed an informed consent prior to the interview. The participants’ anonymity and the confidentiality of the data were ensured. The interview was part of the DaiLiHOT_2 study approved by The Danish Research Ethics Committee (H-22032988) and the Danish Data Protection Agency j.nr. P-2022-625.

## Results

All invited patients accepted to participate, and thus a total of 12 patients, eight men and four women, were interviewed. In five cases the patient’s spouse also participated. Patients’ characteristics are shown in Table 2. The interviews lasted from 18 to 44 minutes.

**Table 2.** Descriptives of the study patients with COPD on LTOT.

| Variables, n=12 |  |
| --- | --- |
| Gender, men/female, no. | 8/4 |
| Age, years | 72.9 ± 5.5 |
| Fixed home oxygen dose, L/min | 1.9 ± 0.8 |
| Time with LTOT, month | 21 ± 17 |
| mMRC, 0-4 | 3.0 ± 0.5 |
| FEV<sub>1</sub>, liter | 0.9 ± 0.2 |
| FEV<sub>1</sub>, % of predicted | 35 ± 12 |
| FEV<sub>1</sub>/FVC, ratio | 0.55 ± 0.44 |
| Marital status, no. (%) |  |
| Lives with spouse | 9 (75) |
| Lives alone | 3 (25) |
| Engages in activities outside their home (self-assessed), no. (%) |  |
| Rarely | 4 (33.3) |
| Regularly | 2 (16.7) |
| Often | 6 (59) |
| Lives in, no. (%) |  |
| House | 4 (33.3) |
| Apartment without lift | 4 (33.3) |
| Apartment with lift or ground floor | 4 (33.3) |

Data are presented as mean ± standard deviation (SD), or counted number with percentage (%). Abbreviations: LTOT: long-term oxygen therapy; mMRC: modified Medical Research Council Dyspnea Scale; FEV<sub>1</sub>: forced expiratory value in the first second; FVC: forced vital capacity.

Our data analysis revealed three themes related to 1) “This is my life”, the life with a chronic pulmonary disease that requires home oxygen therapy for daily living, 2) “Getting the oxygen I need”, the patients’ perception of having their oxygen saturation automatically adjusted, and 3) “New technology gives hope for my life”, the patients experience of having the automated oxygen equipment installed in their home.

The themes and subthemes are presented in Figure 2 and described below, illustrated with selected patients quotes.

**Figure 2.**
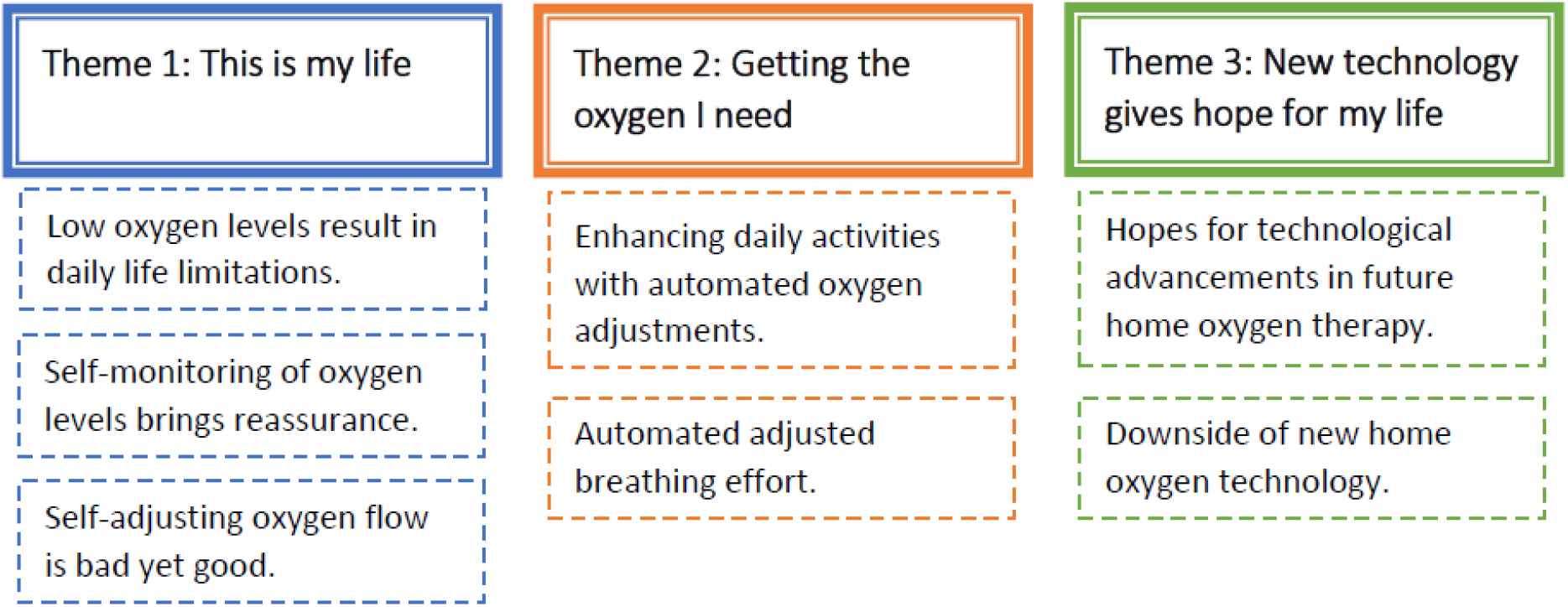
Overview of themes and subthemes

### Theme 1, This is my life

The patients described the emotional, physical, and social burden of living with COPD and relying on oxygen supplement. They expressed feelings of sadness, isolation, and dependency, coupled with an awareness that their condition and physical function would not improve, but rather deteriorate as the disease progressed. In addition, the presence of oxygen equipment was perceived by the patients as a constant reminder of the severity of their illness.

> *“…I realized; this is my life.” – George, in his late 70s.*

### Subtheme: Low oxygen levels result in daily life limitations

The combination of breathlessness, reliance on oxygen, and mental stress created difficult situations for the patients. Some patients expressed how the feeling of running short of oxygen forced them to stop even basic tasks like going to the bathroom. Even while using their usual oxygen therapy, they often found themselves unable to continue with their activities and described this struggle as a direct result of not getting enough oxygen.

> *“It comes automatically, I mean if you have no oxygen, you stop. Like the Duracell bunny. When there’s no more power, you just stop.” – Peter, in his early 60s.*

This need to pause frequently due to a drop in their oxygen saturation caused a range of symptoms and it was described by several of the patients as a regular disruption in their daily routine.

> *“I can feel it when it drops. Because… I get really tired. I feel exhausted and have to sit down. I get… dizzy, simply. And I… I feel unwell. I can literally feel it… like when I was over in… in… where it went wrong. I had to sit down (in the elevator at the supermarket)” – Arthur, in his late 70s.*

Furthermore, some patients reported feelings of anxiety, particularly in situations where they faced heavy breathlessness.

> *“I’m trying to convince myself: You’ve been through this before, and it will pass soon. But sometimes it can be hard to convince oneself.” – Lucas, in his late 70s.*

However, others described how they had found ways to accept their limitations and reconcile themselves to their circumstances, acknowledging breathlessness and the need of oxygen supplement as a part of their illness. They described how they no longer lived in constant fear of running short of oxygen.

> *“It is uncomfortable. I don’t get nervous because I use to say: if it happens, it happens. You can’t do shit about it.” – William, in his late 70s.*

### Subtheme: Self-monitoring of oxygen levels brings reassurance

As a part of daily routines, the patients described how they normally monitored their own oxygen saturation. They had acquired a small finger pulse oximeter, which they used frequently throughout the day and carried with them. The patients described how they used the pulse oximeter as a form of self-assessment to validate the severity of their perceived breathing difficulties, providing tangible evidence of their dyspnea.

> *“Sometimes I say: ‘Oh, now I can feel that I’m lacking oxygen.’ ‘Yeah, yeah,’ my wife says*… *And then I could see it was down to 80. So, I got a good answer to my suspicion or my feeling about how it was*… *You CAN literally feel the difference.” — George, in his late 70s.*

The patients described self-monitoring as a valuable tool for understanding their oxygen levels, which they considered essential. Over time, they learned to sense their saturation through their breathing, consistently linking low oxygen levels to increased breathlessness and discomfort, referring to low saturation as ‘no fun.’

> *“It’s the oxygen saturation that’s decisive again, I mean, when you*… *if you go down to below 70 or down to around 70, right? Well, then*… *then it’s really no fun anymore.” – Peter, in his early 60s.*

### Subtheme: Self-adjusting oxygen flow is bad yet good

The patients explained how physicians and nurses independently had instructed them not to adjust their home oxygen flows. They acknowledged being instructed that increasing the oxygen flow could have safety implications, but the rationale behind the restriction was not of primary concern to the patients. Whether they fully understood or agreed with this aspect of their oxygen therapy was not evident from their statements.

> *“I’ve been told that I’m not allowed to adjust anything. DO NOT touch anything!” – Emma, in her late 70s.*

Despite regularly monitoring their oxygen saturation and recognizing when they were running low on oxygen, the majority of patients emphasized their adherence to these medical instructions, but primarily out of respect for their doctors’ authority rather than personal conviction.

> *“They said to keep this at 1.5, you know, and I don’t want to go against what the doctors said.” - Jack, in his late 60s.*

However, there were also some rebels. A few patients described how they adjusted the oxygen flow upward during heavy physical activities, fully aware that this went against the usual recommendations.

> *“Oh no, you mustn’t do that. No, no, I say, I don’t do that*… *but, well, I do. (laughs) I mean, I have to get up the stairs.” – Julia, in her mid-60s.*

They had been advised not to adjust the oxygen themselves, but the patients explained how they had noticed that ‘more oxygen’ allowed them to engage in tasks that required extra effort, which justified their self-adjustments in the oxygen flow.

> *“If, for example, I need to wash the floor or something, I just turn it up in there… Well, it helps me, so…!” – Lydia, in her mid-70s.*

### Theme 2: Getting the oxygen I need

The patients frequently described automated oxygen therapy by comparing it to their usual treatment with fixed oxygen flow. Their comparisons were rooted in their familiar reactions and experiences with home oxygen, making it challenging for them to express their experiences with the new treatment without referring back to what they already knew.

### Subtheme: Enhancing daily activities with automated oxygen adjustments

Much in line with their interest in monitoring their oxygen saturation with usual oxygen, the patients described using both the wrist pulse oximeter – to assess their oxygen saturation- and the tablet, which additionally showed the amount of oxygen delivered by the automatic device. They reported that their oxygen saturation was more stable during the intervention period, and they also expressed fascination in noticing that their oxygen needs were sometimes lower than what they normally received. They referred to themselves as being curious and captivated by the fluctuations in their oxygen flow and how these changes affected them.

> *“I’m intrigued, you know, how fast I am recovering now” – Jack, in his late 60s.*

The patients expressed confidence in the electronic device’s ability to deliver the precise amount of oxygen they needed. They highlighted a sense of relief and reassurance, knowing that their oxygen flow was automatically adjusted to meet their needs. This was especially confidence-building during physical activities typically associated with discomfort and breathlessness.

> *“I felt safe with it. It helps me; if I use more energy, it also gives me more energy… It somehow gave me peace of mind. I didn’t need to worry about that because it would balance itself out.” – Emma, in her late 70s.*

The patients described how the optimized oxygen flow allowed them to move more naturally, enabling them to perform daily tasks without frequent breaks. They noted greater flexibility in managing their activities, increasing the likelihood of completing tasks within their time limits.

> *“*…*I was able to remove some spider webs*… *I could actually vacuum. I also cleaned the kitchen floors slowly and calmly. Things were much easier. And you weren’t tired in the same way” – Julia, in her mid-60s.*

### Subtheme: Automated adjusted breathing effort

The patients reported experiencing fewer episodes of breathlessness while using the automated oxygen system and they described less mental effort or strain when trying to control their breathlessness. They explained that breathing required less conscious focus or mental energy to manage.

> *“… you didn’t really have to think about what you were doing, right? You just did it.” – William, in his late 70s.*

Additionally, some patients reported a decrease in the intensity of breathlessness. They found that they could perform activities with less breathing effort and with a sense of relief or improved well-being.

> *“… you feel much better. Suddenly, you realize you can move around without gasping and struggling for air.” – Julia, in her mid-60s.*

Some patients felt more refreshed in the mornings, and some described feeling closer to their ‘normal’ selves. Many noted a faster recovery after breaks and told that when they paused to catch their breath, they were able to resume their activities after a much shorter break than usual.

> *“I’ve already said that I miss it… When you do something: you don’t get out of breath as quickly, and you get your pulse down faster. It speaks for itself.” – Peter, in his early 60s.*

### Theme 3: New technology gives hope for a better life

#### Subthemes: Hopes for technological advancements in future home oxygen therapy

The patients shared numerous reflections on the new technology installed in their homes. They described how the system functioned and where they positioned the devices to ensure optimal signal. The patients’ interest in the technology was evident through thoughtful suggestions for improvements, such as a patch that could be discreetly placed on the body, a built-in Bluetooth sensor in a watch, or a portable version of the automated oxygen device for use while away from home.

> *“I think it would be annoying to always have to walk around with a sensor on your finger… If it could be done with a patch that could be applied, I think that would be much simpler… and not annoying at all.” – Andrew, in his late 60s.*

However, some patients also expressed a wonder about why so little progress had been made in the field of home oxygen therapy compared to our otherwise highly digital world, while others remained optimistic, believing that rapid development would occur in the near future.

> *“I am sure once they find out: oh, this is a good thing! … the airplane was discovered in 1902, yes, but by 69 we were on the moon, like the technology goes fast, the development, so I’m very confident they will sort something out…” – Jack, in his late 60s.*

Furthermore, some patients raised concerns about who would cover the costs of these new technologies and if they would be a reality for patient with lung diseases.

#### Subthemes: Downside of new home oxygen technology

The patients consistently reported significant annoyance due to the persistent noise generated by the oxygen concentrator. The patients perceived the continuous humming of the concentrator as particularly bothersome, and they described it as the most substantial challenge associated with automated oxygen therapy.

> *“The biggest problem is the noise. It really is. I had to move it” – Arthur, in his late 70s.*

Consequently, finding sufficient space to place the concentrator in a separate room or as far away as possible from the patients’ immediate living areas became a critical concern.

> *“It (the oxygen concentrator) was out here in the hallway*… *the one I’m using now. The big one, I put in the room, right? Because it was so noisy.” – William, in his late 70s.*

Patients living in small homes described the difficulties they faced, emphasizing that there was simply not enough space to position the concentrator at a comfortable distance. Some patients shared how they had to move the concentrator between the bedroom and living room depending on where they were, while one explained that he chose to sleep on the couch to avoid the noise, leaving the bedroom for the concentrator. A few patients came up with creative solutions, such as placing the concentrator in a broom closet, or using whatever extra space was available, like the bathroom.

The patients further expressed dissatisfaction with the pulse oximeter, which they found too cumbersome and consuming too much battery power.

> *“It is half a grandfather clock.” – Peter, in his early 60s.*

The patients described their experiences with alarms from the equipment and how they responded to them. Their reactions and approaches varied significantly from feeling helpless and startled to calmly reconnecting different components with a sense of control and composure. Some mentioned with a shrug that it was simply a matter of restarting the device. They saw no difficulty in handling such interruptions. In contrast, others recounted situations where they had no idea what to do and had to call for assistance from a family member or their contact person in the study.

> *“I thought I was being clever and could save some electricity. So… foolish as I am, I turned it off. And then Cleo [the automated device] went out, everything went out. And Cleo was just standing there, beeping away. I put her, that Cleo, in my laundry basket… all the way down there. Oh lord, it was so annoying. Beep beep beep” – Ida, in her mid-70s.*

When the patients were asked directly whether they would want the equipment reinstalled as it was at the time - with its noise, the large pulse oximeter with the finger sensor, and the teething problems - 9 out of 12 patients answered, ‘Yes, please’. Some patients justified their responses by explaining that life was easier with automatic oxygen, that the device ‘did something for them,’ or that it simply worked better for their needs. Others mentioned that it was logical to want more oxygen when they felt short of breath. A few expressed disappointments that such a device was not yet available, and one patient humorously attempted to offer pastries to the interviewer in exchange for a device. A small number of patients quickly mentioned their primary wishes for improvements.

> *“Yeah, I would jump on it without hesitation. Yeah.” – Jack, in his late 60s.*

A few patients declared that they did not feel any different with the automated oxygen at all.

## Discussion

To our knowledge, the 12 patients in this study were the first to experience second-by-second oxygen adjustments at home based on their oxygen saturation. The patients expressed a desire to maintain stable oxygen saturation within the target range, as they associated hypoxemia with dyspnea, discomfort, and difficulties with performing activities of daily living. They reported to believe, that the automated oxygen titration supplied them with the oxygen that they needed and that this gave them a feeling of less dyspnea and was useful when performing everyday tasks. The patients were very interested in the new technology and positive about its installation in their homes. They also offered suggestions for improvement, primarily focused on reducing the noise from the concentrator.

### Patient perception of oxygen status in the body

Our results illustrated how the patients were highly interested in and attentive to their oxygen saturation levels. This demonstrates the importance the patients placed on the oxygen aspect of their condition. For instance, all but one patient already owned a pulse oximeter before enrolling in the study, having obtained it on their own, and routinely measured their oxygen saturation as part of their daily life. The interest in self-monitoring is not unfamiliar. A scoping review with 16 studies, including various patient groups, found that both patients, healthy individuals, and healthcare professionals were generally positive about the use of pulse oximeters, even on a daily basis [17]. The patients expressed a clear desire for an immediate oxygen flow response following their needs. Conversely, some patients found it interesting to observe that they received less oxygen while resting or sleeping. These findings, emphasize the need for continuous and dynamic oxygen adjustments to meet patients’ varying needs throughout the day, rather than for instance making infrequent, small changes.

The sensation of dyspnea during exercise in patients with COPD can arise from various factors, including increased work of breathing, hypoxemia, or physiological factors like anxiety [18]. It is possible that patients find it difficult to differentiate between these contributing factors during activity, as their dyspnea could result from a combination of them. However, hypoxemia is a known source of dyspnea and will undoubtedly contribute to this sensation [19]. The patients in the present study consistently expressed that desaturation affected them and described the sensation as unpleasant. This unpleasant dyspnea may also explain why self-monitoring became so important to the patients, as it provided a way to manage their condition.

If the patients felt how lack of oxygen affected them negatively it would prompt them to avoid or minimize the duration of hypoxemic episodes and force them to stop or even avoid certain tasks altogether. This is exactly what the patients described occurring when performing activities of daily living. Conversely, the patients noted that they experienced changes in their breathing during physical activities when their oxygen flow was being adjusted, enabling them to do their daily routine with less dyspnea and a faster recovery. Moreover, some patients reported a sense of safety or reassurance with automated oxygen titration, which for some could be related to their perception that their oxygen saturation was more stable. This narrative aligns well with other quantitative studies examining the effect of optimal oxygenation during physical exercise, showing that an improved oxygen saturation can be clinical meaningful to the patients with an improved physical performance and breathing in patients with COPD [10,11,14,20].

In a qualitative study by Goldbart et al. examining patients’ experiences with LTOT compared to not receiving supplemental oxygen, patients reported several benefits, including increased confidence, faster recovery, and improved performance and breathing [21]. The patients in our study fundamentally expressed the same experiences of improved oxygenation through automated oxygen titration as the patients in Goldbart’s study. The trustworthiness of the patients’ experiences is further reinforced by the previous mentioned study, in which optimized oxygenation during a semi-lab ADL-test increased the patients’ ability to perform ADL and alleviated their feeling of dyspnea [14].

The patients’ narratives also reflected an increased ability to perform physically demanding activities with less cognitive effort. Some patients reported that receiving the right amount of oxygen allowed them to simply “be,” as it relieved a sense of mental fog. Since hypoxemia can lead to cognitive impairment and reduced mental clarity [4,22], this could explain why the patients felt more like themselves when their exposure to hypoxemia was minimized. Patients with COPD often describe their breathing as requiring mental effort. During daily activities, a group of patients in a study by O’Donnell et al. predominantly selected the terms “air hunger” and “my breathing requires mental effort or concentration” in the Multidimensional Dyspnea Profile (MDP) questionnaire [23]. In a randomized controlled trial, Sandau et al. found that patients hospitalized for acute exacerbation reported a high perception of mental breathing effort, also measured by the MDP [9]. Sandau et al. used automated oxygen titration to optimize oxygen levels, resulting in a reduction of dyspnea and a significant decrease in the sensation of mental effort. In summary, mental breathing effort is a common experience for patients with COPD, and it seems like reducing hypoxemia could alleviate this burden. This highlights the significance of our patients’ statements when they expressed, “I did not have to think, I just did it.”

The patients reported that they were not permitted to adjust the oxygen flow on their usual care concentrators at home. This restriction may be due to the risk of hypercapnia if patients either forget to readjust the flow, or if they make uninformed decisions about their oxygen dose. Such situations could have severe consequences. Additionally, there is currently no evidence showing that minimizing periods of hypoxemia affects patients’ well-being, or disease prognosis. However, as previously mentioned, several studies demonstrate a significant effect of improving oxygenation on perceived dyspnea and performance during exercise, activities of daily living, and dyspnea during acute exacerbations in COPD [9-11,14]. These positive effects on patient-reported outcomes, combined with the feasibility of automated oxygen titration at home and the strong interest in optimal oxygenation from the patients in the present study, provide a solid foundation for further research.

### The technology

“We are sending rockets to the moon,” reflected a patient, thereby highlighting the contrast between advancing technology and the current state of supplemental oxygen therapy. While the oxygen concentrator was not directly part of the automated equipment, an automated system still requires, not only a closed-loop device, but also a large, stable concentrator, a pulse oximeter with a strong Bluetooth signal, and optimal coordination between these elements. Nearly all patients voiced complaints about the noise generated by the concentrator, which was a standard 9 L model. Particularly when the concentrator was positioned closer to them, it affected their comfort. A further challenge, according to the patients were the continuous adjustment by the pulse oximeters. These devices need a strong Bluetooth connection to prevent significant signal loss, especially if the patient lives in a large house. As the patients noted, the wrist pulse oximeter was bulky and inconvenient, requiring a battery change every other day. One patient suggested creating a hidden patch, which the authors find to be an interesting idea for the future.

Installing the automated oxygen equipment in the patients’ homes involved more than simply adding a new appliance. This device was responsible for managing the oxygen therapy, including determining the appropriate flow rate, and timing of delivery. This transition naturally captured the patients’ attention. Interestingly, while patient preferences regarding the development of oxygen equipment have not been extensively researched—and were not a primary focus of our study - their engagement with their oxygen therapy and their feedback on the technology generated insightful reflections. Their reaction to the device alarms varied significantly, which likely reflected different levels of experience, willingness, and ability to address technical challenges. These perspectives suggest that it is important to secure a minimum level of technical understanding so that the patient can react adequately to an alarm. Listening to the patients’ concerns and being aware of user-friendly automated systems and quieter concentrators could drive innovation and improve the quality of home-based oxygen therapy.

The statement from a patient that development would probably be possible if someone were willing to put money into “patients with COPD” reflects a feeling of being marginalized or undervalued. This sentiment could be interpreted as an expression of health inequity, as we see a group of patients who must endure constant noise from life-sustaining concentrators and intermittent hypoxia, yet often lack the energy, skills, or organizational support to advocate for themselves. As the need for advancements in home oxygen therapy becomes increasingly apparent, there is hope that interest in developing such technologies will grow. A recent ERS statement calls for urgent development on oxygen systems, which we can only applaud [24]. We hope for technologies that will allow patients to receive up to 8 liters of oxygen from a quiet flow-strong concentrator based on their saturation, using a compact, rechargeable pulse oximeter that does not take up the space of “half a grandfather clock.”

### Strength and limitations

Given that the first author was involved in the DaiLiHOT_2 study, the potential for allegiance bias cannot be entirely dismissed. Although we sought to mitigate this risk by selecting an interviewer not involved in previous studies to conduct most of the interviews, this possibility still remains. Additionally, the patients were not blinded to the intervention, which could have influenced them to view automated oxygen titration more positively, as they could observe how their oxygen saturation became more stable.

The willingness of the patients to participate in the DaiLiHOT_2 study and accept the equipment in their homes may suggest that they represent a more open-minded group toward automated oxygen therapy, which could limit the generalizability of the findings to a broader COPD population. While the sample size might be considered limited, it is important to recognize that in qualitative research, a group of 12 participants is typically considered adequate [25]. However, this number could still be seen as small for broader representation.

A notable strength of this study was the successful recruitment of 100% of eligible participants, despite their severe lung disease and limited energy for extended conversations. These patients were more than willing to share their experiences, providing valuable insights. We also believe it is a strength that the interviews were conducted in the patients’ homes with an interviewer not wearing a uniform, creating an informal atmosphere. This setting, along with the participation of patients’ spouses, encouraged honest and open responses.

The data we gathered was rich and comprehensive, and by the time the final interview was conducted, we had reached a point of information power, with no new themes emerging, which strengthened the reliability of our analysis. Furthermore, to ensure the trustworthiness of our results, multiple authors were actively engaged in the analysis process. Themes and subthemes were discussed iteratively, and the interdisciplinary nature of the author group, with affiliations to different organizations, helped minimize bias and avoid preconceived assumptions from influencing the analysis and interpretation.

## Conclusion

In this study, the patients with COPD on long-term oxygen therapy valued maintaining their oxygen saturation within a target range, as they associated hypoxemic episodes with unpleasant dyspnea. The patients reported improved ability to engage in daily activities when their oxygen levels were adjusted automatically based on their saturation using automated oxygen titration. Incorporating patient feedback into the design of future oxygen delivery systems could enhance both implementation and patient satisfaction. Further research over a longer duration is needed to evaluate potential long-term benefits of automated oxygen titration.

A special thanks to all the patients for sharing their valuable perspectives.

## Data Availability

All data produced in the present study are available upon reasonable request to the authors

